# Prediction of the coronavirus epidemic prevalence in quarantine conditions based on an approximate calculation model

**DOI:** 10.1101/2020.05.17.20104810

**Authors:** Denis Below, Felix G. Mairanowski

**Affiliations:** M.Sc. Psychology; Professor, Doctor of Science

## Abstract

A calculation model for predicting the spread of the COVID-19 epidemic under quarantine conditions is proposed. The obtained simple analytical ratios allow estimating the factors determining the intensity of the infection spread, including changing requirements for quarantine severity. The presented method of forecasting allows to calculate both the total number of infected persons and the number of active infections. Comparison of the results of calculations according to the proposed model with the statistics for a number of cities shows their satisfactory qualitative and quantitative compliance. The proposed simple model can be useful in preliminary assessment of possible consequences of changing quarantine conditions.

## Model description

Currently, mainly models based on the numerical solution of the system of ordinary differential equations [1, 2] are used to calculate the spread of the Coronavirus epidemic. The numerous coefficients of these models are selected by comparing the results of calculations with observations of the spread of the epidemic found in different countries or individual cities. Under conditions of various measures, including isolation of disease carriers by quarantine or other sanitary measures, there is a need to assess the effectiveness of these restrictive measures. Thus, it becomes vital to develop a simple model, which allows to quickly assess the effectiveness of the strategy to combat the uncontrolled spread of the epidemic. It should be understood that the modeling of the spread of the epidemic should be carried out under conditions of low reliability of information on the number of infected persons, due to the fact, in particular, that the results of observations depend to a large extent on the quantity and quality of the tests for the presence of the disease.

The simplest class of models for the spread of the epidemic are those based on the analysis of the growth of the population of infected persons. The basic equation of the growth rate of infected persons in a closed control zone (city, country) is shown:

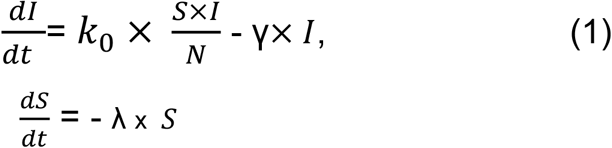

In which:

*I = the number of infected persons at a given time*,
*k*_0_ *= Coronavirus infection rate (1/day)*
*N = total population of the area under consideration*,
*S =* the number of susceptibles, *a part of the population that has the potential to become infected through contact with infected persons*,
γ = *the rate of treatment and mortality of coronavirus patients, i.e. the rate of transition of persons from the category of infected to the category of persons immune to infection*.

In this case, it is assumed that under quarantine conditions, the intensity of disease is artificially restrained by reducing the contacts of infected persons with persons who potentially can get infected by the ratio obtained from the solution of the second equation of the system (1):

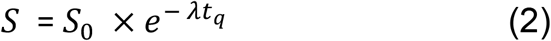

In which:

*S*_0_ *= maximum number of potentially infectious persons*,
*λ = intensity factor of decrease in contacts of infected patients with with persons who potentially can get infected by means of quarantine and other preventive measures*.
*t_q_ = current time from quarantine initiation*.

This equation assumes that under quarantine conditions, parameter S depends not on the number of infected persons I, but on quarantine stiffness conditions, i.e. λ is a function of 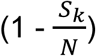, where *S_k_* is the number of people who potentially can get infected but are removed from active contact with infected patients when quarantined.

Thus, the first original model equation takes shape:

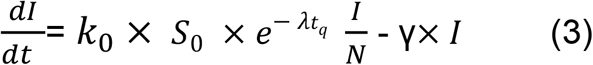

The solution to this equation is:

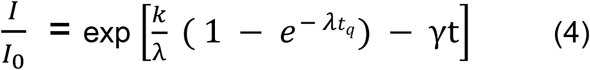

In which *I*_0_ is the number of initial infection.

This ratio is the basic equation for calculating the spread of the virus coefficient.

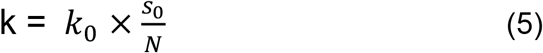

The initial number of infected patients can be estimated by the ratio obtained from the condition that at the beginning of the epidemic it develops according to a simple exponential growth.

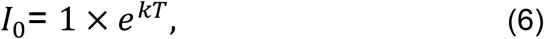

*T = period of time per day from the beginning of detection of the first patients until quarantine is introduced*.

Coefficient k can be estimated according to (5) as a product of two parameters. The maximum intensity factor according to observations in the USA and Spain [3] can be defined as the minimum period of doubling the number of cases t_2min_ = 1 day. Then *k*_0_ = ln(2)= 0,691/day. The second part of the equation in (5) can be estimated if we take into account that, according to preliminary data from virologists at the Clinic Charite in Berlin [4], about 34% of the population not in contact with carriers of coronavirus may have resistance to this disease. The second multiplier in the ratio (5) can therefore be estimated as 0.65, and therefore k=0.69 × 0.65 = 0.43 1/day. It can be assumed that this factor ranges from 0.4 to 0.6 per day. For larger cities where the intensity of contacts is higher than for cities with small populations, this factor should generally be higher as the number of contacts increases with population growth. It is possible to determine this factor more precisely from the statistics for the period from the beginning of the epidemic to the introduction of quarantine from the formula (6):

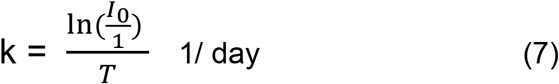

(Here 1 is entered for dimensions).

However, the accuracy of the calculation in (7) is currently limited by the lack of reliability of information of the number of infected patients at the beginning of the epidemic.

The coefficient λ in (4) determines the effectiveness of measures to reduce the intensity of virus transmission both as a result of reduced quarantine contacts and the use of various preventive sanitary and hygienic measures. Since quarantine is the most extreme means to combat the spread of the epidemic, we will link this coefficient to the amount of people of those left who potentially can get infected after the quarantine to their amount without being in quarantine. For this purpose, we will introduce the concept of maximum infection when quarantine of this severity is introduced without considering the effect of treatment, i.e. at γ = 0 and at t → ∞:

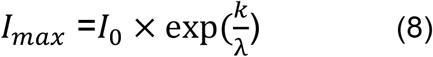

When the strictness of the quarantine changes, the number of infected persons changes *I_max_* and, in turn, when the specified value is *I_max_*, the coefficient of λ is calculated as

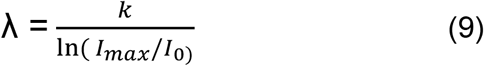

## Examples of calculations

The coefficient γ of reduction I as a result of recovery of patients is determined exclusively by the results of statistics and depends on the level of development of health care, availability of free places in hospitals, qualification of medical personnel, sanitary and hygienic conditions, and environmental conditions. Let us first analyze the total number of patients infected with the virus for the entire period of the epidemic without taking into account the cure factor at γ = 0.

The model coefficients k and λ for most cities, including Berlin and New York, were calculated on the basis of statistical data presented in [6] and [2], for Moscow - in accordance with the data presented in [5]; the coefficient k was calculated on the basis of the ratio (7) for Berlin k = 0.4, for Moscow - 0.5, for New York - 0.54. The parameter λ was assumed to be the same and equal to 0.13 for Berlin and New York, since the level of strictness of the quarantine in these cities was approximately the same and for Moscow it was slightly lower (0.12). The analysis of the epidemic spread in large cities revealed that for example in Madrid, the quarantine severity was higher than in Berlin, so λ = 0.15 was assumed there. In example, Fig. 1 shows the results of the proposed model calculations for Berlin, New York and Moscow.

**Fig. 1:**
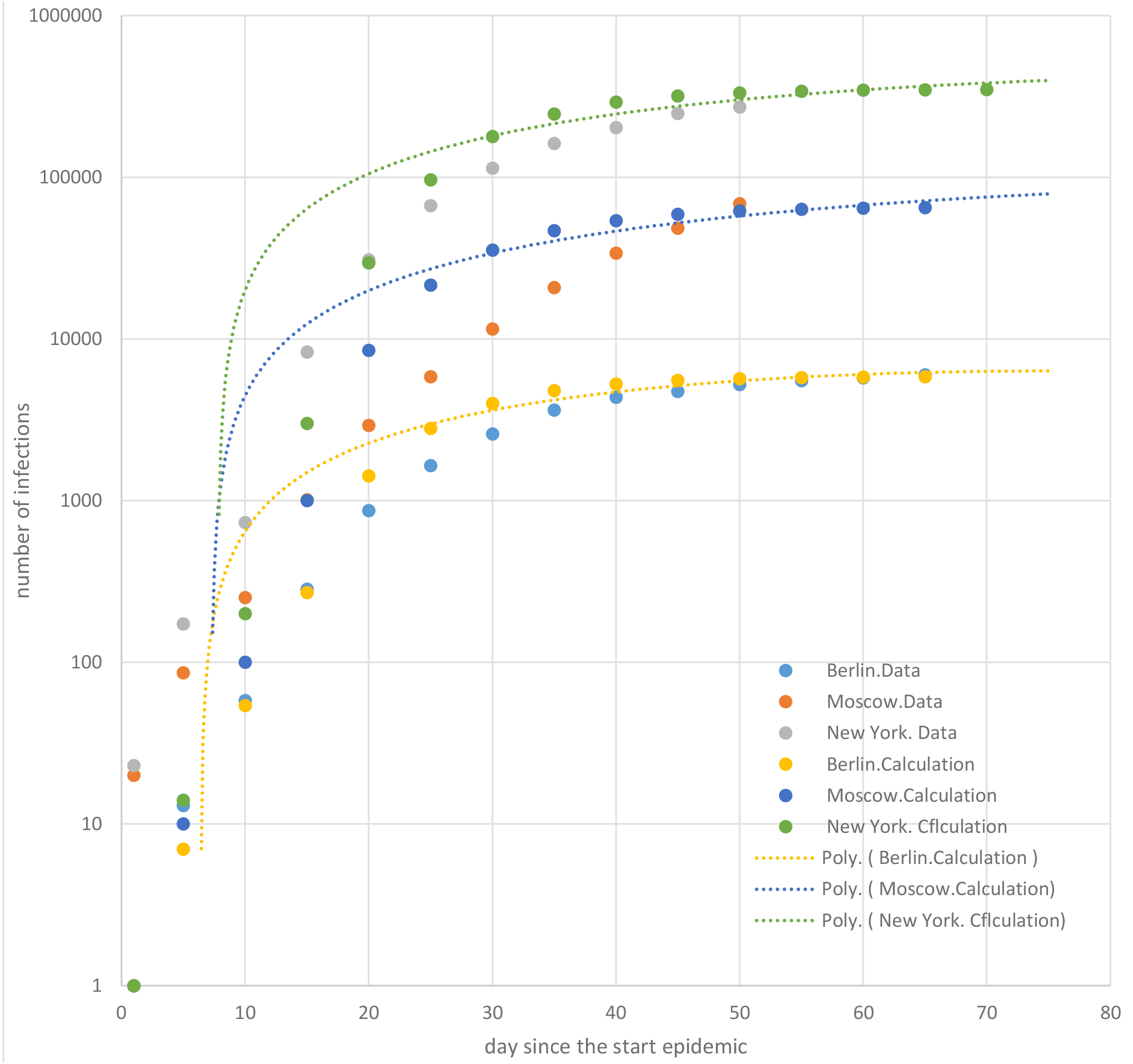
The spread of the coronavirus epidemic cumulative infection

The comparison of the calculation results with statistical data shows satisfactory qualitative agreement. As for quantitative correspondence, it is explained by the choice of model coefficients. It can be noted that statistics on the number of infected persons for different countries is extremely unreliable, as it depends on the volume of testing. This can be seen especially well from the analysis of data for Moscow [5], where the number of established infections has increased sharply only about 30 to 40 days after the official beginning of the epidemic. According to official information, during this period the number of tests in Moscow doubled. The number of tests is particularly low and therefore there was a low level of infection with the virus detected in the early stages of the epidemic. As reliable information on the spread of the epidemic accumulate, the choice of model coefficients will become more and more reasonable. The unreliability of the information became apparent, in particular, in the calculation of the coefficient k for Moscow; this coefficient should have been about 0.54 (i.e., the same as for New York) and, accordingly, the number of infected persons, according to the predicted calculations, would have increased to about 120000.

With the successive removal of the number of quarantine restrictions, an assessment of the possible consequences of such measures is necessary. Fig. 2 shows the results of the forecast of possible virus spread in Berlin for two infection scenarios: *I_max_* = 200,000 and *I_max_* = 400,000 (i.e. about 5% and 10% of the Berlin population). In both cases, according to the calculation, the epidemic would continue for about a year after the quarantine was removed. The active phase of the epidemic will continue for approximately 8 months.

**Fig. 2:**
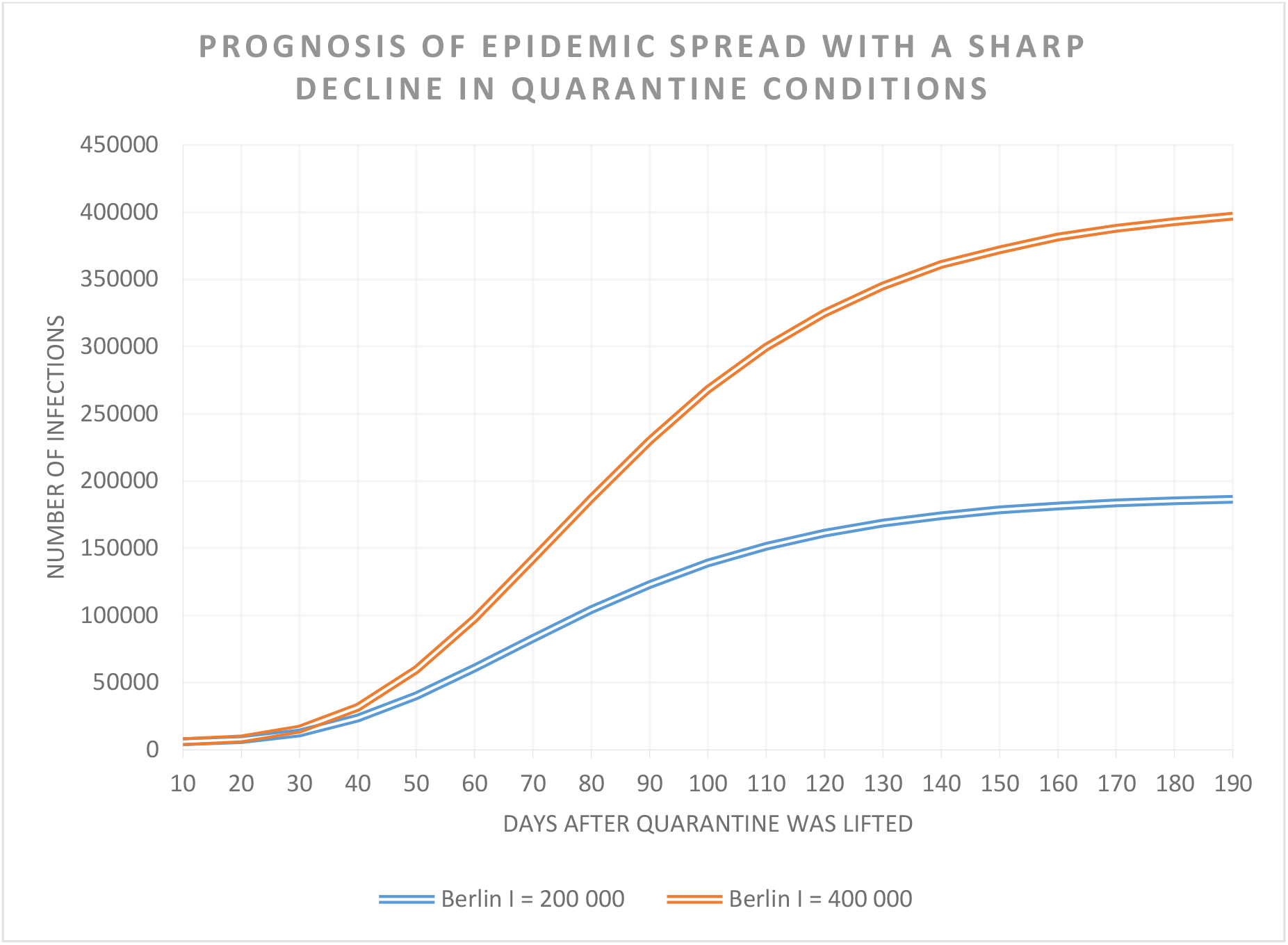
Prognosis of epidemic spread with a sharp decline in quarantine conditions

The predicted increase in the number of infested persons after easing of quarantine conditions or quarantine withdrawal was also calculated with a similar ratio (4). The time in this case is counted from the moment the quarantine conditions change and the total number of infested persons is determined as the sum of them during the quarantine period and after the quarantine requirements decrease.

Having received the second derivative from the ratio (4) (at γ = 0) in time and having equated it to zero, it is possible to define the time, counted from the beginning of quarantine conditions easing, at which the maximum growth of infected patients will be observed.

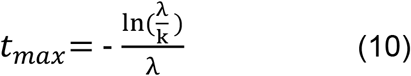

For the calculated cases under consideration, the maximum daily increase in patients infected with the virus is more than 5000 persons for *I_max_* = 400 000 and more than 2000 persons for *I_max_* = 200 000.

The scenario of a slower easing of quarantine requirements was also considered. The proposed model allows estimating the rate of change in the number of infected patients over time depending on the quarantine level. Thus, for example, if for Berlin the number of infected patients increases by Δ*I* when quarantine severity decreases, then we can estimate the new values of λ by (9) with the replacement of *I_max_* byΔ*I*.

The use of the prognostic model makes it possible to determine the maximum possible easing of quarantine conditions (maximum number of potentially infectious cases) that will not lead to an excessive overload of the city’s health care system. The results of the prognostic calculations show that for the Berlin context, the maximum daily increase in the number of infected patients is about 40 days after the quarantine attenuation. If the number of infected patients can be expected to double compared to the number of patients under strict quarantine, the daily maximum increase in the number of infected patients will reach over 150 per day. It is calculated that a new stationary regimen (i.e. a total of about 14000 infected patients) could be expected after about four months. However, after three months it would be possible to produce a secondary quarantine easing. In case of quarantine weakening, a more successful quarantine strategy could be used, leaving mainly the persons in the so-called risk zone in the hard quarantine. For this purpose, first of all, it would be necessary to weaken the quarantine for residents who are not in the so-called risk group. However, it is necessary to bear in mind that the risk group may include not only the elderly, but also young citizens and even children with a certain set of diseases. But such a strategy will require mass testing and analysis of the health status of a large number of citizens and, most importantly, a reliable determination of the number of persons with resistance to disease. In addition to analyzing the total number of people infected with the virus, information on the number of active patients, i.e. actually infected patients, is equally important, taking into account their cure. The calculation of these so-called active patients was performed on the basis of a ratio (4).

Fig. 3 shows the total number and the number of actively infected patients for Berlin based on the results of statistical data and obtained by the proposed model calculation. The cure coefficient γ was taken equal to 0.03. Analysis of this graph shows that the number of actively infected citizens in Berlin *I_a_* begins to decrease after 35 - 40 days from the beginning of the virus epidemic.

**Fig. 3:**
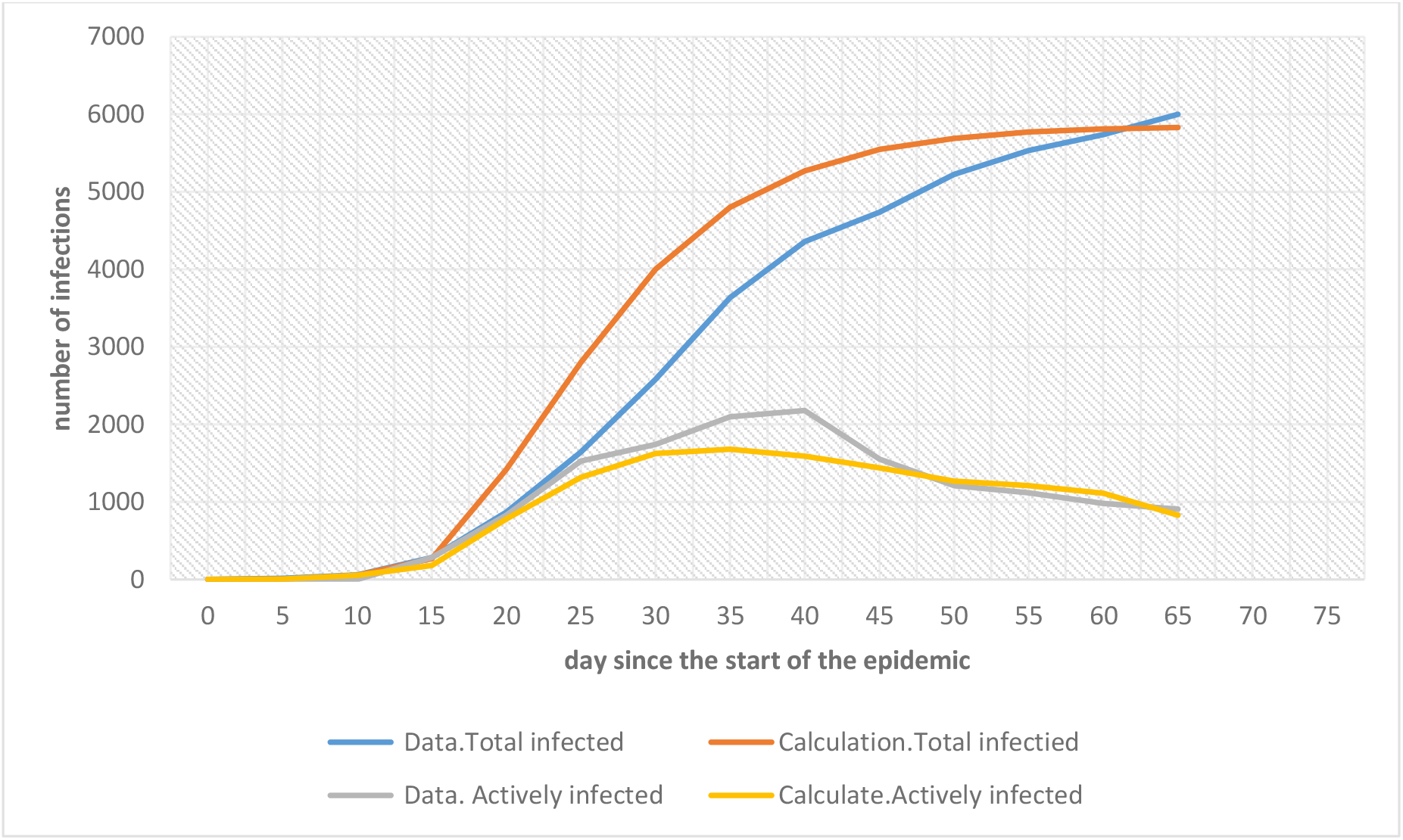
The spread of the epidemic in quarantine conditions

The period of time after which it can be expected that the number of active infections begins to decrease is, when the so-called *R*_0_ ≤ 1 can be determined from the ratio (4). The infection rate *R*_0_ is the parameter that controls the number of new infections in a certain moment given the number of infected people in the days before. For *R*_0_ ≤ 1 the infection curve becomes flat and the epidemics is controlled. This condition corresponds to a simple condition of (4): 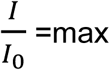. By equating the first derivative (4) to zero after simple transformations we get:

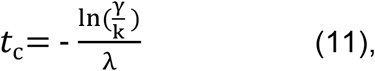

In which

*t*_c_ *= time, counted from the beginning of quarantine conditions easing, after which the epidemic becomes controlled (R*_0_ *≤ 1)*.

In general, the results of the model calculations are qualitatively satisfactory with statistical data. Better quantitative correspondence could have been easily achieved by adjusting model coefficients, but this was not the task of this paper. It was more important to show that a simple analytical solution makes it possible to assess the effectiveness of measures aimed at containing the virus epidemic in a quite satisfactory way.

The calculation of active infections makes it possible to make an alternative prediction of the change in the number of infected patients after the quarantine conditions have changed. This calculation is also performed by the ratio (4), but in formula (9), *I_a_* is taken instead of *I*_0_ when determining the coefficient λ.

Result of such an alternative forecast calculation for Berlin for the values *I_max_* 400000 and 200000 and *I_a_* = 800 allowed in particular to establish that the maximum daily increase in the number of infected persons occurs significantly earlier than it is presented in Fig. 2. So, for example, for conditions *I_max_* = 400 000, in about a month after the quarantine weakening, the daily maximum growth of infected persons reaches almost 7000 people per day.

The choice of methods for calculating the change in the number of infected persons after a radical weakening of quarantine conditions will be based on comparing the results of the predicted calculations for both methods with statistical data. The first method is based on the assumption that by the time of quarantine weakening the epidemic is almost over, i.e. the development of the epidemic begins again. With the second method of calculation, it was assumed that by the time quarantine weakened, the epidemic may develop again intensively due to contact with actively infected persons. But both methods of calculation show the danger of almost uncontrolled epidemic development in case of an abrupt change in quarantine conditions.

## Conclusions and direction for further research

1. A simple model has been developed to quickly forecast the spread of the COVID-19 epidemic under quarantine and other preventive measures aimed at drastically reducing the number of infections.
2. Comparison of results of calculations by the proposed model with statistical data showed satisfactory qualitative and quantitative agreements.
3. The model uses two main empirical parameters determining the intensity of the epidemic spread: the contact intensity coefficient k determining the population infection dynamics and the contact restriction or infection effect reduction coefficient as a result of different sanitary and epidemiological measures λ.
4. The use of this model as a tool for further numerical experiments allows to estimate the timelines and effectiveness of measures to reduce the spread of the epidemic. For example, the model can be used to calculate the number of infected citizens in New York City under the hypothetical condition that quarantine would have been introduced earlier than it happened in reality.
5. The most important factor determining the further quarantine exit strategy is the information about the potential number of persons who have already received immunity against the virus. Reliable information can only be obtained through mass testing of the population. There is currently no critical information on the number of people who may have developed immunity to the virus, including as a result of their already asymptomatic or very mild coronavirus disease. According to some preliminary data, the number of such persons who were immune after asymptomatic or mild disease may be 5-10 times greater than the number of statistically recorded infections, therefore the relative resistance to coronavirus may reach 70-80% [7].
6. Further specification of the spread of the infection can also be done with the help of analysis of the impact of the disease on different individual groups of the population. At the same time, it would be useful to use a more detailed model with separate predictions for different population groups and the possibility to forecast the number of severe diseases and the death rate.
7. The use of the proposed model makes it possible to quickly assess the consequences of planned measures for partial or complete removal of quarantine restrictions or introduction of new restrictions.
8. Further improvement of the model, first of all, should be carried out in the direction of clarifying the coefficient k and determining the change of its values depending on it:
  a. The value of the locality for which the forecast is made: Preliminary data obtained by us show that the value of this coefficient increases with population growth.
  b. Population groups: these groups can be divided by age, social origin, and ethnic composition. There is abundant evidence of varying degrees of epidemic prevalence among children, middle-aged people and the elderly. Statistics also show an increased risk of disease for weaker social groups, which may be due to both higher rates of violations of certain health standards in these groups and more serious health problems.
  c. Vaccination rates in the population: Some data show that vaccination against other viruses, which was mandatory, particularly in Eastern European countries, may have helped to reduce the spread of coronavirus COVID – 19.
  d. Climatic characteristics: Preliminary studies carried out at the Massachusetts Institute of Technology (MIT) [9] show, inter alia, that with increasing temperatures and decreasing absolute humidity, the intensity of the epidemic decreases.
  e. Special attention should be paid to analysing the impact of long-term epidemics on the psychological state of the population: It should be noted that even a relatively short period of the epidemic (about 3 months) has sharply increased the risk of stress and depression [8]. Therefore, when choosing an optimal strategy to combat the epidemic, one should also take into account the possible worsening of mental diseases and the impact of long-term quarantine on them. In this regard, it would be essential to perform an analysis of the risk of developing mental illness both in conditions of long-term quarantine and active epidemic development without the use of severe quarantine restrictions.
  f. This work was carried out in the initial stage of the epidemic development. As a result, it was impossible to establish the main parameters of the model reliably enough. Only after quarantine it will be possible to provide a refined analysis of the model’s coefficients and, possibly, determine in more detail the directions for further research.

## Data Availability

[1] Fred Brauer, P. van den Driessche, and Jianhong Wu. (2008). Lecture Notes in
Mathematical Epidemiology. www.researchgate.net/publication/265887931.
[2] Noll, N. B., Askamentov, I., Druelle, V., Badenhorst, A., Jefferies, G., Albert, J., &
Neher, R. (2020). COVID-19 Scenarios: an interactive tool to explore the spread and
associated morbidity and mortality of SARS-CoV-2. medRxiv.
www.medrxiv.org/content/10.1101/2020.05.05.20091363v2.full.pdf.;https://covid19-
scenarios.org
[3] Fagen-Ulmschneider W. (2020). An interactive visualization of the exponential
spread of COVID-19. www.91-divoc.com/pages/covid-visualization/ (Zugriff
02.05.2020)
[4] Drosten C, Henning K. (2020). Coronavirus Update Folge 16.
www.ndr.de/nachrichten/info/coronaskript184.pdf (Zugriff 02.05.2020)
[5] Coronavirus distribution map in Russia and worldwide. (Zugriff 02.05.2020).
Yandex. www.yandex.ru/maps/covid19
[6] Der Regierende Bürgermeister von Berlin - Senatskanzlei (2020). Fallstatistik
Coronavirus. www.berlin.de/corona/fallstatistik (Zugriff 02.05.2020)
[7] Hoffmann, D. (2020). Hohe Dunkelziffer: Die ersten Ergebnisse der Con-VinceStudie. Luxemburger Wort. www.wort.lu/de/lokales/hohe-dunkelziffer-die-erstenergebnisse-der-con-vince-studie-5eb42a8eda2cc1784e35d406.
[8] Quervain, D., Aerni, A., Amini, E., Bentz, D., Coynel, D., Gerhards, C., Zuber, P.
(2020). The Swiss Corona Stress Study. https://doi.org/10.31219/osf.io/jqw6a.
[9] Bukhari, Q., & Jameel, Y. (2020). Will coronavirus pandemic diminish by
summer?.

## References

[1] Fred Brauer, P. van den Driessche, and Jianhong Wu. (2008). Lecture Notes in Mathematical Epidemiology. www.researchgate.net/publication/265887931.

[2] Noll, N. B., Askamentov, I., Druelle, V., Badenhorst, A., Jefferies, G., Albert, J., & Neher, R. (2020). COVID-19 Scenarios: an interactive tool to explore the spread and associated morbidity and mortality of SARS-CoV-2. *medRxiv*. www.medrxiv.org/content/10.1101/2020.05.05.20091363v2.full.pdf.;https://covid19-scenarios.org

[3] Fagen-Ulmschneider W. (2020). An interactive visualization of the exponential spread of COVID-19. www.91-divoc.com/pages/covid-visualization/ (Zugriff 02.05.2020)

[4] Drosten C, Henning K. (2020). Coronavirus Update Folge 16. www.ndr.de/nachrichten/info/coronaskript184.pdf (Zugriff 02.05.2020)

[5] Coronavirus distribution map in Russia and worldwide. (Zugriff 02.05.2020). Yandex. www.yandex.ru/maps/covid19

[6] Der Regierende Bürgermeister von Berlin—Senatskanzlei (2020). Fallstatistik Coronavirus. www.berlin.de/corona/fallstatistik (Zugriff 02.05.2020)

[7] Hoffmann, D. (2020). Hohe Dunkelziffer: Die ersten Ergebnisse der Con-Vince-Studie. Luxemburger Wort. www.wort.lu/de/lokales/hohe-dunkelziffer-die-ersten-ergebnisse-der-con-vince-studie-5eb42a8eda2cc1784e35d406.

[8] Quervain, D., Aerni, A., Amini, E., Bentz, D., Coynel, D., Gerhards, C., Zuber, P. (2020). The Swiss Corona Stress Study. https://doi.org/10.31219/osf.io/jqw6a.

[9] Bukhari, Q., & Jameel, Y. (2020). Will coronavirus pandemic diminish by summer?

